# Factors that affect enrollment into a community-based health insurance scheme among households in Arba Minch Zuria District, Gamo Zone, Southern Ethiopia, 2023: A community-based unmatched case-control Study

**DOI:** 10.1101/2024.03.04.24303762

**Authors:** Silas Bukuno Bujitie, Yusuf Haji daud

## Abstract

**Background:** Many low-and middle-income countries are affected by catastrophic health expenditures as out-of-pocket payments exceed the World Health Organization’s (WHO) 10% threshold level. The government of Ethiopia has been working to reduce out-of-pocket payments from 37% to less than 15% in its health sector transformation plan II; however, the scheme was marked by a low membership and dropout rate. The aim of the study was to identify determinants of enrolment into a community-based health insurance scheme (CBHIS) in Arba Minch Zuria District of Gamo Zone, Southern Ethiopia.

**Methods:** Unmatched case-control study was employed from February 8, 2023, to March 9, 2023, among 327 participants (109 cases and 218 controls to community-based health insurance (CBHI) scheme) in a 1:2 proportions. Multi-stage sampling technique was used to draw the study participants. Data were collected by using structured interviewer administered questionnaire and then entered into Epi-data and exported to SPSS for analysis. Bivariable and multivariable analysis was carried out using binary logistic regression. Significance was declared by using an adjusted odds ratio (AOR) with 95% confidence interval (CIs) and a p value of <0.05.

**Results:** A total of 327 participants (109 enrolled and 218 non-enrolled) were interviewed, resulting in a response rate of 100%. Family size (AOR=1.66; 95% CI: 1.00–2.73), wealth index (AOR=2.29; 95% CI: 1.25–4.19), awareness level of the community based health insurance scheme (AOR=3.78; 95% CI: 1.09–13.18), and perceived quality of health care (AOR=1.67; 95% CI: 1.02-2.75) were found to be determinant factors of enrollment in the scheme.

**Conclusion:** Strengthening community awareness activities, focusing on families of poor households, and improving the quality of health service delivery are highly recommended to improve enrollment in the scheme.

## Introduction

Community-Based Health Insurance (CBHI) is an instrument that increases access to quality healthcare for communities working in the informal sector and offers financial protection against the cost of disease. The other type of health insurance is social health insurance (SHI), which includes people who work in the formal economy[1]. The 2030 Sustainable Development Goals (SDGs) place a premium on ensuring that all people have access to high-quality healthcare without financial burden[2]. Access to adequate health services is a prerequisite for people to achieve good health and lead a healthy lifestyle[3]. Universal Health Coverage (UHC) ensures that all individuals and communities have access to high-quality health care without financial burden. Every year, catastrophic health expenses push hundreds of millions of people into poverty in low-and middle-income countries (LMICs)[4]. Many LMICs faced challenges in raising adequate funds to finance health care for their citizens[5]. Thus, they are encouraging risk-pooling institutions, like CBHI, to finance healthcare out-of-pocket payments (OOPs)[6].

CBHI is a community-based strategy administered by the government or a private, for-profit organization that provides insurance services to cover the cost of health care[7]. Since the 1990s, many LMICs use it as a strategy to avert OOPs for purchasing health services[8]. The CHBI scheme is a risk-pooling approach that shares the financial burden among households with varying health statuses to make it more accessible[9]. Globally, over 150 million people face financial hardship because of healthcare expenditures; as a result, the number of households in deep poverty increases by about 25 million households each year[10]. The 2005 WHO assembly achieved UHC by implementing health insurance programs and reducing direct payments out of pocket. Global healthcare spending is less than 12% of total Gross Domestic Product (GDP)[4, 11].

Globally, over 800 million people spend 10% of their income on health care through direct payment, potentially increasing the number of people entering poverty each year[12].

Because of limited resources, many African households fund their healthcare costs by selling personal property[13]. More than half of Sub-Saharan countries have higher direct healthcare spending, which is greater than 30% of total income[14]. 42% demonstrated this in Kenya[15] and 37% in Ethiopia[16]. The high amount of direct payments for health care in Ethiopia hinders access to health services[17]. The Ethiopian government spends less than 5% of its GDP on health, which is less than the requirement set by the Alma Ata Declaration, which is 15%[18]. The Ethiopian government aims to reduce OOPs to less than 15% in Transformation Plan II[19]. Community-based health insurance is an alternative approach for countries such as Ethiopia that cannot directly cover the costs of medical care for the poor[20, 21]. Thus, Ethiopia launched a pilot CBHI scheme in June 2011 in 13 different districts of Amhara, Oromia, SNNPR, and Tigray regional states[22], with the future intent to cover 83.6% of the population of Ethiopia who are engaged in informal sectors, mostly rural residents[23]. According to the Ethiopian Health Insurance Agency’s (EHIA) CBHI report, it has reached 50%; however, the 2020 target was to achieve 80% coverage[24]. This number decreases to 42% in the study district, with varying performance across its kebeles in 2021/2022[25]. Although it has shown progress since its inception, the scheme has been marked by low membership and a high dropout rate from the program once members have joined[6, 26]. Low membership is associated with low enrollment at the outset or a high dropout rate after joining the scheme[27]. There was no study that documented the determinants of enrollment in CBHI in Gamo Zone in general and Arba Minch Zuria district in particular. This study aimed to figure out the determinants of enrollment in the CBHI scheme in the Arba Minch Zuria District.

## Materials and Methods

### Study Setting and Period

The study was conducted from February 8, 2023, to March 9, 2023, in the Arba Minch Zuria District of Gamo Zone, Southern Ethiopia. Arba Minch town is the administrative town of the district, which is located 275 km from the regional capital Hawassa and 505 km from Addis Ababa (the country’s capital). Arba Minch Zuria District is one of the 20 districts of Gamo Zone. Administratively, the district is divided into one urban and 18 rural kebeles. According to the 2007 national census projection, the estimated total population of the district was 129,665, in 2022, of which 49 percent were male. The district has 26 health centers and 4 health posts, which are under the government, and 32 clinics owned by the private sector. Primary health care coverage in the district was 80%. According to the district CBHI scheme department, there were about 26,462 households in the district, of which 23,552 (89%) were eligible for the CBHI scheme. The rest, 11%, were expected to be engaged in the formal sector. According to the department’s 2021–2022 annual report, 9,891 (42% of eligible) households were enrolled in the scheme. Among enrolled households, 2,410 (24%) were needy[25]. Fig 1 shows administrative map of the study area.

**Fig 1:**
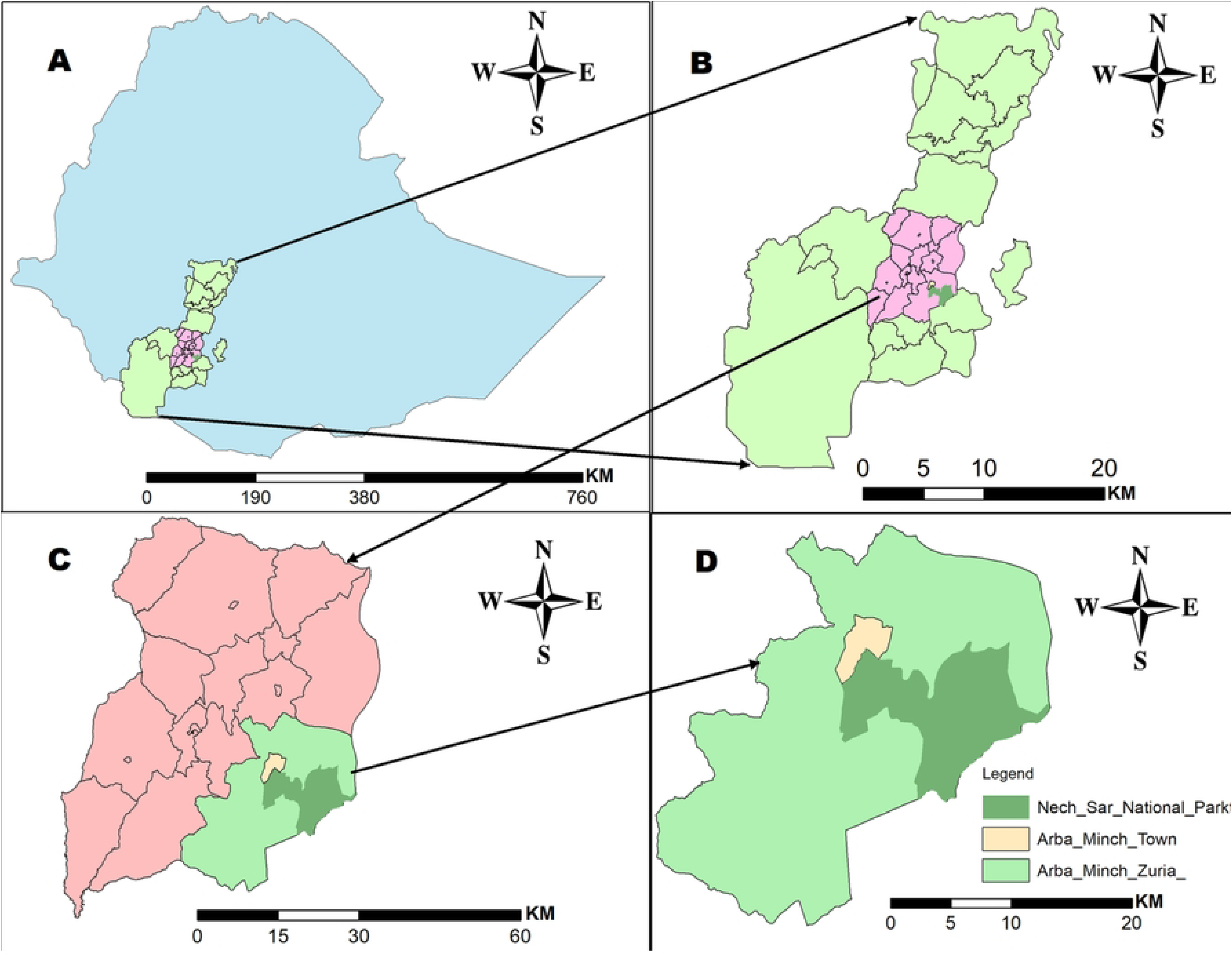
(As indicated in fig 1, **A**-represents the Ethiopian boundary, **B**-represents the South Nation Nationalities and Peoples’ (SNNP) Regional State, **C**-represents the Gamo Zone, and **D**-represents the administrative map of Arba Minch Zuria District)

### Study Design

An unmatched community-based case-control study was used.

### Populations

**Source population:** all households that resided in Arba Minch Zuria District during data collection were source populations.

**Study population:** all randomly selected households in selected kebeles of the district were considered as a study population.

## Eligibility criteria

### Case

**Inclusion criteria:** residents who lived in selected kebeles for at least six months and were formally registered for the scheme for the years 2021-2022 during data collection were included in the study.

**Exclusion criteria:** household heads who worked in the formal sector or had health insurance through another scheme and were unable to communicate (critically sick) during data collection were excluded from the study.

### Control

**Inclusion criteria:** non-enrolled households that lived for at least six months in the selected kebeles of the district during the study period were included in the study.

**Exclusion Criteria:** non-enrolled household heads in any form of insurance scheme who were unable to communicate (Critically ill) during the study period were excluded from the study.

## Sample size determination and sampling procedure

### Sample size determination

The double population proportion assumption formula was used to calculate the sample size using Epi-Info software version 7. The following parameters were considered: a precision of 5% at a 95% confidence level and a power of 80%. The ratio of controls to cases (r) = 2, OR = 2.79, P1 = 29.5%, and P2 = 17.2% [28]. In addition, a 10% possible non-response rate was multiplied by the design effect of 1.5. Finally, a sample size of 327 (109 cases and 218 controls) was obtained.

### Sampling procedure

A multi-stage sampling technique was used to select samples. In the first phase, six kebeles were selected by simple random sampling (the lottery method) from a pool of 19 kebeles, considering their representativeness. In the later stage, we used the same technique to select households from six selected kebeles. A list of households in the selected kebeles was collected from the health posts. The sample size for selected kebeles was determined by the number of households in each kebele (Proportional allocation). Figure 2 illustrates a diagrammatic presentation of the sampling procedure.

**Fig 2:**
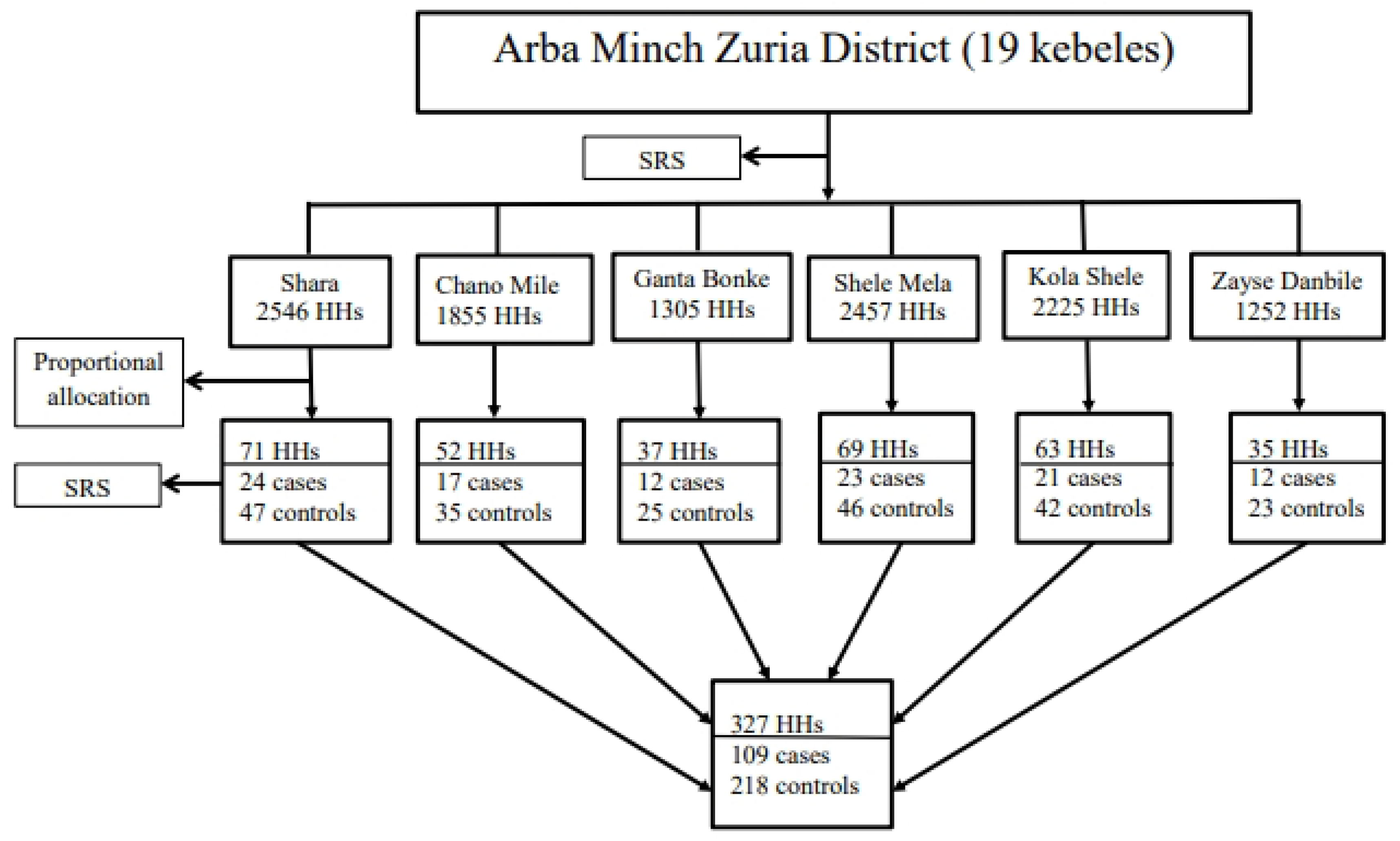
Schematic presentation of sampling procedure, Arba Minch Zuria District, Gamo Zone, Southern Ethiopia, 2023

## Study variables

### Dependent variable

CBHI enrollment status

### Independent variables

The independent predictors were classified as socio-demographic factors (age, sex, marital status of household heads, household size, educational level, occupational status, and family wealth index), individual-related factors (awareness level of CBHI, perception towards CBHI, and perceived quality of health services), household-related factors (chronic or recent illness in the household and elderly people in the household), facility-related factors (accessibility of health facilities and availability of health services), and scheme-related factors (premium affordability and benefits package)

### Operational definitions

***Wealth index:*** this variable was evaluated using 30 household assets: live stocks, household infrastructure (electricity, bicycle, electric bakery, bed, mattress, and stove), latrine, water source, housing condition (floor, wall, roof, window, kitchen, separate cattle room), and size of agricultural land. After dichotomizing all thirty variables into yes/no or improved/unimproved categories, principal component analysis was used to reduce the dimensionality of large data sets. Finally, the wealth index was categorized as poor, medium, and rich for analysis in SPSS.

***Awareness about the CBHI scheme:*** participants’ awareness of CBHI was assessed using five questions that comprised its existence, its principles, and the significance of community-based health insurance. Participants who correctly responded to this question were classified as “correct responses, while those who did not were classified as “incorrect responses.” Each item has an equal value. Thus, the correct response scored 1, and the wrong response scored 0. Finally, the aggregate score for all awareness questions ranges from 0 to 5 points. Participants’ overall awareness was categorized as good if the scores were 3 and above (>= 60%) and otherwise poor.

***Perception towards the CBHI:*** this is how people perceive the CBHI’s benefits package adequacy, service quality; trust in the management body, and patient handling by health professionals. This is measured via five-point Likert scale questionnaires ranging from “strongly disagree’ to “strongly agree,” which are used for respondents to express their opinions on four perception tools. To measure this variable, first, we computed the median of the individual sample, then the sample median score was calculated and graded as positive perception (for values above the median score) and negative perception (for values less than the median score). The Cronbach’s alpha result for this variable was 0.91.

***Perceived healthcare quality:*** this was the extent of the respondent’s view on the quality of healthcare delivery. This includes the health worker’s perspective, the availability of drugs and supplies, including diagnostics, the waiting time, and the cleanliness of the facility. This is measured via five-point Likert scale questionnaires ranging from “strongly disagree’ to “strongly agree,” which are used for respondents to express their opinions on five perception tools. To measure this variable, first, we computed the median of the individual sample, then the sample median score was calculated and graded as good quality (for values above the median score) and poor quality (for values less than the median score). The Cronbach’s alpha result for this variable was 0.79.

***Affordability of premium:*** affordability was used in this research to assess the perception of household heads on premium affordability (375 ETBR per year): whether it is low, medium, or expensive.

***Chronic illness:*** a disease condition that lasts over 3 months.

### Data collection tools and techniques

We used a structured questionnaire adapted from the Ethiopian Demographic and Health Survey (EDHS) to collect data[29]. Originally, the questionnaire was prepared in English, and it was also restated into Amharic and back to English by a language expert to ensure consistency. The questionnaire contains socio-demographic characteristics, individual-related factors, household-related factors, facility-related factors, and scheme-related factors. Three BSc nurses and two public health officers were involved in data collection and supervision independently. They were trained for one day to become familiar with data collection tools. Health professionals were named based on their previous experience in data collection. Supervisors check the completeness and clarity daily and send a report for data entry.

### Data quality management

One-day training was given for each data collector and supervisor, focusing especially on the privacy and confidentiality of the study participants. Supervisors check the data daily for completeness and accuracy before data entry. The data collection tool was pre-tested at 5% of the sample size in the non-selected kebele of the study district.

### Data processing and analysis

The data were cleaned and edited after being entered into Epi-data version 4.6.0.6 and exported to Statistical Package for the Social Sciences (SPSS) version 27 for analysis. All categorical variables were cross-tabulated with response variables and presented using frequency, proportions, and tables for case and control groups. All explanatory variables that were significantly associated with the response variable in the bivariable logistic regression analysis at p<0.25 were entered into the multivariable logistic regression analysis using the backward elimination stepwise method to identify independent predictors in the CBHI enrollment. The model’s acceptability was checked using the Hosmer and Lemeshow goodness-of-fit test and the Omnibus test. Adjusted odds ratios (AORs) with their corresponding 95% confidence intervals (CIs) were used to claim the significance of predictor variables at a P-value of 0.05.

### Ethical considerations

Ethical clearance was obtained from the institutional ethics review board (IRB) of Hawassa University under the reference number IRB/105/14 and the study was conducted in accordance with Declaration of Helsinki for human subjects. Written informed consent was also obtained from participants before data collection. They were informed that participation will be voluntary. The right to withdraw from the study during the interview was assured. No personal identifiers were used in the data collection tool. No data were accessed by a third party except the principal investigator and were confidential.

## Results

### Socio-demographic characteristics

All three hundred twenty-seven participants (109 CBHI users and 218 non-CBHI users) got interviewed, resulting in an overall response rate of 100%. Most cases (79%) and controls (80%) were male. The mean age of cases and controls was 38.9 years (SD=+ 10.2), and 39.4 years (SD=±11). Nearly forty-six percent (45.9%) of participants were found under the age category of 36–54 years for both cases and controls. Regarding marriage, 92.7% of cases and 89% of controls were married. Nearly 47% of cases and 59% of controls were farmers (Table 1). The Bivariable logistic regression analysis of CBHI-program related variables were also described in (Table 2).

**Table 1:** Bivariable logistic regression analysis of socio-demographic variables for CBHI enrolment in Arbaminch Zuria District, Gamo Zone, Southern Ethiopia, 2023.

| Explanatory Variables |  | CBHI Enrollment Status |  | COR | 95% CI |  | P-value |
| --- | --- | --- | --- | --- | --- | --- | --- |
|  |  | Enrolled (N/%) | Not Enrolled (N/%) |  | Lower | Upper |  |
| Age | 18-35 | 45(32.8) | 92(67.2) | 1 |  |  |  |
|  | 36-54 | 50(33.3) | 100(66.7) | 0.98 | 0.59 | 1.60 | 0.930 |
|  | ≥55 | 14(35) | 26(65) | 0.91 | 0.43 | 1.91 | 0.799 |
| Sex | Male | 86(33) | 175(67) | 1 |  |  |  |
|  | Female | 23(34.8) | 43(65.2) | 0.92 | 0.52 | 1.62 | 0.770 |
| Ethnicity | Gamo | 79(35.1) | 146(64.9) | 0.85 | 0.46 | 1.59 | 0.616 |
|  | Wolaita | 12(26.7) | 33(73.3) | 1.27 | 0.53 | 3.02 | 0.589 |
|  | Others<sup>a</sup> | 18(38.3) | 39(68.4) | 1 |  |  |  |
| Religion | Protestant | 66(32.2) | 139(67.8) | 1.81 | 0.58 | 5.58 | 0.305 |
|  | Orthodox | 37(33.9) | 72(66.1) | 1.67 | 0.52 | 5.32 | 0.387 |
|  | Others<sup>b</sup> | 6(46.2) | 7(53.8) | 1 |  |  |  |
| Marital Status | Single | 3(33.3) | 6(66.7) | 1 |  |  |  |
|  | Married | 101(34.2) | 194(65.8) | 0.96 | 0.24 | 3.92 | 0.955 |
|  | Others<sup>c</sup> | 5(21.7) | 18(78.3) | 1.80 | 0.33 | 9.89 | 0.499 |

**Table 1:-** (Continued).
|  |  |  |  |  |  |  |  |
| --- | --- | --- | --- | --- | --- | --- | --- |
| **Family size** | 1-5 | 70(40) | 105(60) | 1 |  |  |  |
|  | >5 | 39(25.7) | 113(74.3) | 1.93 | 1.20 | 3.10 | 0.006* |
| **Educational status** | No education | 18(34) | 35(66) | 1 |  |  |  |
|  | Primary (1-8) | 56(32.7) | 115(67.3) | 1.05 | 0.55 | 2.02 | 0.870 |
|  | 2<sup>nd</sup>ry & above | 35(34) | 68(66) | 0.99 | 0.49 | 2.01 | 0.998 |
| **Occupation** | Farmer | 51(28.5) | 128(71.5) | 2.26 | 1.22 | 4.17 | 0.009* |
|  | Merchant | 31(34.1) | 60(65.9) | 1.74 | 0.89 | 3.43 | 0.108 |
|  | Daily laborer | 27(47.4) | 30(52.6) | 1 |  |  |  |
| **Wealth index** | Poor | 45(38.1) | 73(61.9) | 1 |  |  |  |
|  | Middle | 40(40) | 60(60) | 0.93 | 0.54 | 1.59 | 0.779 |
|  | Rich | 24(22) | 85(78) | 2.18 | 1.22 | 3.92 | 0.009* |
| <sup>a</sup>Zayse & Goffa, <sup>b</sup>Muslim & Atheist, <sup>c</sup>Divorced & widowed 1=Reference variable, *Variables significant at p-value 0.25<br>COR-Crude odds ratio |  |  |  |  |  |  |  |

**Table 2:** Bivariable logistic regression analysis of CBHI-program related variables for enrolment in Arbaminch Zuria District, Gamo Zone, Southern Ethiopia, 2023.

| Explanatory Variables | CBHI Enrollment Status |  | COR | 95% CI |  | P-value |
| --- | --- | --- | --- | --- | --- | --- |
|  | Enrolled (N/%) | Not Enrolled (N/%) |  | Lower | Upper |  |
| Presence of elderly people (≥ 60 years) |  |  |  |  |  |  |
| Yes | 20(37.7) | 33(62.3) | 0.79 | 0.43 | 1.46 | 0.458 |
| No | 89(32.5) | 185(67.5) | 1 |  |  |  |
| Presence of chronic patients |  |  |  |  |  |  |
| Yes | 6(30) | 14(70) | 1.18 | 0.44 | 3.15 | 0.744 |
| No | 103(33.6) | 204(66.4) | 1 |  |  |  |
| Awareness level of CBHI |  |  |  |  |  |  |
| Poor awareness | 106(35.6) | 192(64.4) | 1 |  |  |  |
| Good awareness | 3(10.3) | 26(89.7) | 4.79 | 1.42 | 16.18 | 0.012* |
| Perception towards CBHI |  |  |  |  |  |  |
| Negative perception | 60(40.3) | 89(59.7) | 1 |  |  |  |
| Positive perception | 49(27.5) | 129(72.5) | 1.78 | 1.12 | 2.82 | 0.015* |
| Perceived health care quality |  |  |  |  |  |  |
| Poor quality | 61(40.9) | 88(59.1) | 1 |  |  |  |
| Good quality | 48(27) | 130(73) | 1.88 | 1.18 | 2.99 | 0.008* |

**Table 2:-** (Continued).
| Explanatory Variables |  | CBHI Enrollment Status |  | COR | 95% CI |  | P-value |
| --- | --- | --- | --- | --- | --- | --- | --- |
|  |  | Enrolled (N/%) | Not Enrolled (N/%) |  | Lower | Upper |  |
| Premium affordability | Expensive | 25(36.8) | 43(63.2) | 1 |  |  |  |
|  | Medium | 46(33.8) | 90(66.2) | 1.14 | 0.62 | 2.08 | 0.678 |
|  | Low | 38(30.9) | 85(69.1) | 1.30 | 0.69 | 2.43 | 0.409 |
| Distance from health facility | <2km | 42(35.3) | 77(64.7) | 0.87 | 0.54 | 1.42 | 0.585 |
|  | 2-5km | 6(31.6) | 13(68.4) | 1.03 | 0.37 | 2.85 | 0.951 |
|  | >5km | 61(32.3) | 128(67.7) | 1 |  |  |  |
| Availability of diagnostics | Available | 66(31.6) | 143(68.4) | 1.24 | 0.77 | 1.99 | 0.371 |
|  | Not available | 43(36.4) | 75(63.6) | 1 |  |  |  |
| Prescribed drugs | Available | 63(39.9) | 95(60.1) | 0.56 | 0.35 | 0.89 | 0.016* |
|  | Not available | 46(27.2) | 123(72.8) | 1 |  |  |  |
| Patient's waiting time | <1 hrs. | 27(32.9) | 55(67.1) | 0.91 | 0.51 | 1.64 | 0.753 |
|  | 1-2 hrs. | 40(36.7) | 69(63.3) | 0.77 | 0.45 | 1.31 | 0.338 |
|  | >2 hrs. | 42(30.9) | 94(69.1) | 1 |  |  |  |
| 1=Reference variable, *Variables significant at p-value 0.25 COR-Crude odds ratio |  |  |  |  |  |  |  |

### Determinant factors for CBHI enrolment

Among twenty variables, family size, respondent’s occupation, household wealth, awareness level of CBHI, perception towards CBHI, perceived health care quality, and availability of prescribed drugs in the contractual facility were significant at a p-value of <0.25 with a 95% CI. The above seven candidate variables were further fitted in the multivariable analysis using the backward likelihood ratio (LR) to control confounders and determine the effect of all predictor variables on the likelihood of CBHI enrollment. In the final multivariable analysis, the impact of determinant factors on household heads’ decision to enroll in the CBHI scheme was assessed. Hence, the multivariable analysis showed households with a family size of over 5 were 1.66 times (AOR = 1.66; 95% CI: 1.01–2.73) more likely to be enrolled in the CBHI scheme compared to a smaller family size. Households with the highest wealth index (rich) were 2.29 times (AOR = 2.29; 95% CI: 1.25–4.19) more likely to be enrolled in the CBHI scheme compared to their counterparts (poor). Household heads who perceived the quality of health care as good were 1.67 times (AOR = 1.67; 95% CI: 1.02-2.75) more likely to enroll in the CBHI scheme than those households who perceived it as poor (Table 3). Finally, the odds of CBHI enrollment were 3.78 times higher among respondents with good awareness (AOR = 3.78; 95% CI: 1.09–13.18) compared to those with a poor awareness level.

**Table 3:**
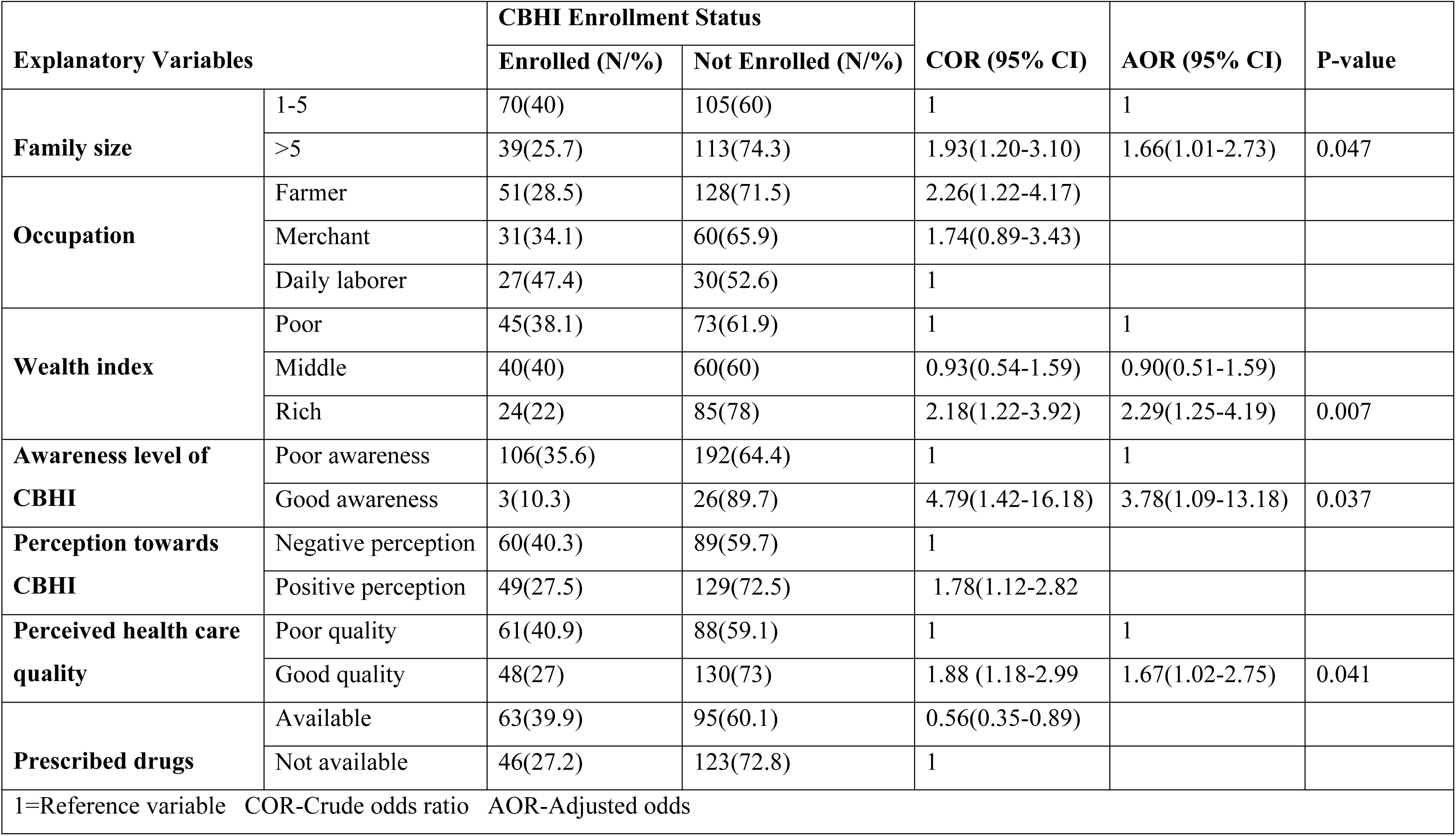
Determinant factors in multivariable logistic regression analysis for CBHI enrollment in Arba Minch Zuria District, Gamo Zone, Southern Ethiopia, 2023.

## Discussions

This research was conducted to identify determinants of the enrollment of households in the CBHI in the Arbaminch Zuria District. Accordingly, the size of the family in the household, the wealth index of the household, the level of awareness of the CBHI and the perceived quality of health care by the household were found to be independent predictors of enrolment in the CBHI scheme.

Our study revealed that households with a larger family size were more likely to enroll in the CBHI scheme than their counterparts. This finding was supported by studies conducted in Ethiopia and Nigeria[30, 31]. The main reason for this could be because the likelihood of having health problems in a family depends on household size. As a result, the likelihood of at least one family member falling ill rises with family size[32]. Therefore, large families have a greater tendency to prefer risk pooling institutions to cover their regular health expenses rather than paying out of pocket[33]. The other reason might be due to the fact that households with a small family size may have a better income and can afford the cost of medical care in better private health facilities[34].

On the contrary, the studies conducted in Burkina Faso[35–37] stands in opposition to our study findings. The possible justification for this might be amount of premium contribution paid by households for membership in an insurance package. In Burkina Faso, the membership contribution for CBHI scheme was estimated per individuals in household, which demands large amount of money for households with large family members. This system might discourage households with large size, as it creates financial burden on the families. Whereas in Ethiopia, the premium contribution was fixed per household level (flat rate), regardless of family size[38]. Another factor that determined the enrollment of households in to CBHI was economic status.

In addition, our study found that households classified as rich by asset category using principal component analysis were twice as likely to enrol in the CBHI programme as those in the poor category. This study’s findings were consistent with a study conducted in Burkina Faso, where enrolment in the CBHI programme is higher among people with higher socioeconomic status[39].

Similar studies in Ethiopia, Nigeria, and India found that wealthier households (the richest quintile) were more likely to enlist in the CBHI scheme than their counterparts[1, 38, 40]. The possible justification for this similarity might be households in the rich category were able to make payment than poor households and there may also be differences in the access to media in the study area. Studies in Uganda and Burkina Faso showed that the primary reason for not-enrolled households were financial problem[39, 41]. Previous research in southwest, western, and southern Ethiopia contradicts our findings that impoverished household heads were more likely to enlist in the CBHI scheme than rich household heads [30, 38, 42]. The justification for this discrepancy might be due to differences in socioeconomic status and health insurance experiences.

Furthermore, enrolment in the CBHI programme is determined by perceptions of health-care quality. Household heads who perceived the health care as of good quality were 1.67 times more likely to enroll in the CBHI scheme than those who perceived it negatively. A similar case-control study conducted in Gida Ayana District, east Wollega Zone, western Ethiopia, confirmed that respondents who perceived high quality of care were found to be 8.37 times more likely to enroll in the CBHI scheme membership compared to those who perceived low quality of care[31]. Another empirical study conducted in Burkina Faso, south Ethiopia and northwest Ethiopia showed consistent evidence with our study findings [36, 42, 43]. This may be justified by the fact that good-quality health services may attract more people. Similar logic works for the CBHI program: the higher society’s perception of the quality of health services, the more people will enroll in the scheme.

Finally, the current study revealed that respondents who had a good awareness level of the CBHI scheme were nearly four times more likely to be enrolled in CBHI compared to those with a poor awareness level. The finding is consistent with a study conducted in Ethiopia[31], where those respondents who had a good awareness level of CBHI scheme membership were 3.78 times more likely to be enrolled in CBHI compared to those with a poor awareness level. Another study conducted in Ethiopia, Bangladesh, and India supported our findings, suggesting that household heads with a good level of awareness of the CBHI scheme were more likely to join the program than their counterparts[44–46]. The possible justification may be due to the fact that people with better information and understanding of CBHI’s existence, principles, and importance are more likely to join the scheme. This implies that raising community awareness may help implementation by improving enrollment in the scheme.

### Limitation of the study

As recall bias was common during the case-control study, however, it was minimised by shortening the time to less than 3 months.

## Conclusion

Households with large family sizes, a high wealth index, a high level of awareness of the CBHI, and perceived quality of health care were found to be independent predictors of enrollment in the study area. Strengthening CBHI awareness activities, focusing on families of poor households, and improving the quality of health service delivery are highly recommended. Finally, to explore other determinants of enrollment in the scheme, researchers should consider mixed-methods research, which may cover a broader geographic area.

## Data Availability

This data will be available to the journal in any form as per the journal's policy.

## Abbreviations and acronyms

CBHI: Community Based Health Insurance
EDHS: Ethiopian Demographic and Health Survey
EHIA: Ethiopian Health Insurance Agency
GDP: Gross Domestic Product
LMICs: Low and Middle Income Countries
SDGs: Sustainable Development Goals
SHI: Social Health Insurance
SNNPR: South Nation’s, Nationalities and Peoples Region
SPSS: Statistical Package for Social Sciences
OOPs: Out-Of-Pocket Payments
UHC: Universal Health Coverage

## Acknowledgements

First and foremost, I would like to thank God Almighty for providing me with the strength, knowledge, talent, guidance, and opportunity to complete my studies successfully. Next, I would like to thank the Ethiopian Field Epidemiology and Laboratory Training Programme and Hawassa University for creating an opportunity to study field epidemiology.

Furthermore, I would like to convey my heartfelt appreciation to my principal adviser, Mr. Yusuf Haji, for his unwavering support and insightful suggestions in the preparation of this thesis. My appreciation also extends to my beloved spouse for encouraging me throughout my study period through prayer. Finally, my thanks go to the data collectors and study participants, without whom this finding would not have been possible.

## Authors’ contributions

**Conceptualization:** Silas Bukuno Bujitie

**Data curation:** Silas Bukuno Bujitie, Yusuf Haji Daud

**Formal analysis:** Silas Bukuno Bujitie, Yusuf Haji Daud

**Funding acquisition:** Silas Bukuno Bujitie, Yusuf Haji Daud

**Investigation:** Silas Bukuno Bujitie, Yusuf Haji Daud.

**Methodology:** Silas Bukuno Bujitie, Yusuf Haji Daud

**Project administration:** Silas Bukuno Bujitie

**Software:** Silas Bukuno Bujitie, Yusuf Haji Daud

**Supervision:** Yusuf Haji Daud

**Validation:** Silas Bukuno Bujitie, Yusuf Haji Daud

**Visualization:** Silas Bukuno Bujitie, Yusuf Haji Daud

**Writing – original draft**: Silas Bukuno Bujitie

**Writing – review & editing:** Silas Bukuno Bujitie, Yusuf Haji Daud

## Funding

There is no funding to report.

## Disclosure

The authors report no conflicts of interest in this work.

## Notes

### Competing Interest Statement

The authors have declared no competing interest.

### Clinical Trial

“N/A”

### Funding Statement

There is no funding for this work.

### Author Declarations

The IRB at the College of Health Sciences of Hawassa University has approved that it complies with all principles for person,beneficence and justice,it is ethically achievable and sound.

